# Elastomeric Harnesses Decrease Aerosol Particles Leak and Improve the Fit of Surgical Masks

**DOI:** 10.1101/2021.08.31.21262838

**Authors:** Jeannette Ingabire, Hannah McKenney, Charles Sebesta, Krishna Badhiwala, Ben Avants, Caleb Kemere, Sahil Kapur, Jacob T. Robinson

**Affiliations:** Department of Electrical and Computer Engineering, Rice University Brown School of Engineering, Texas 77030; Division of Surgery, The University of Texas MD Anderson Cancer Center, Houston, Texas 77030; Department of Plastic Surgery, The University of Texas MD Anderson Cancer Center, Houston, Texas 77030

## Abstract

**Importance:** The outbreak of Coronavirus diseases 2019 (COVID-19) disease has increased demand for N95 respirators, surgical masks, and other facial coverings to stop the spread of SARS-CoV-2. Research shows that N95 respirators perform the best at filtering viral droplets and aerosols, however these masks are much more difficult to manufacture and expensive to distribute on a large scale, which led to shortages during the pandemic. Surgical masks, on the other hand, were more widely available and have been previously used to mitigate the spread of tuberculosis and influenza.

**Objectives:** To evaluate the filter filtration efficiency (FFE) of three elastomeric harness designs in hospital and research settings in order to improve facemask seal.

**Design, setting and participants:** A multi-institutional collaboration between engineers and health professionals, conducted between November 2020 and March 2021, was set up to design an elastomeric harness to improve the face seal of a surgical mask. Three elastomeric harness designs were created with harness designs 1 and 2 tested in a research laboratory setting and harness design 2.1 tested in a hospital setting. The initial harness design 1 was laser cut for testing and design 2 was developed to improve the detected particle leakage around the nose bridge area by introducing more material in that region. Design 2.1 is developed for hospital settings with less material around the nose bridge to reduce vision disruption. The designs were tested on mannequins and human volunteers using IR imaging and standard fit testing equipment.

**Main Outcomes and Measures:** Our elastomeric harness can improve the seal of a surgical mask allowing it to pass the fit test used to evaluate N95 respirators. 24/39 participants achieved a passing score of 100 or more while wearing the second harness design. IR imaging determined that the nasal sidewalls region of the mask is most prone to leakage when using our first elastomeric harness.

**Conclusions and Relevance:** Overall, these results confirm that elastomeric harnesses combined with surgical masks improve their ability to filter aerosolized particles, which is especially important when in close proximity to individuals who are infectious or while performing aerosol-generating medical procedures.

## INTRODUCTION

The emerging and highly contagious coronavirus disease 2019 (COVID-19) global pandemic, caused by the coronavirus 2 (SARS-CoV-2), has increased demand for N95 respirators, surgical masks, and a variety of other facial coverings.^1^ In addition to face masks, public health institutions recommend physical distancing to decrease the spread of SARS-CoV-2 and flatten the infection curve.^1,2^ SARS-CoV-2 affects the respiratory tract, with higher viral concentration in respiratory secretions and saliva.^3,4,10^ Research shows that communities that implement mask-wearing are associated with reduced COVID-19 prevalence as a result of reduced viral droplet exposure from infected individuals.^5^ Meta-analysis work show that wearing face masks reduce the transmission of respiratory illnesses such as COVID-19.^6^

Although it is scientifically clear that face masks alleviate the spread of COVID-19 on a population level, knowledge on the efficiency associated with different types of face masks is still limited. A recent study that evaluates optical measurements of droplet transmission shows that while medical grade masks such as N95 respirators and surgical masks perform the best at preventing droplet transmission, commonly used fabric ones like neck-gaiters and bandanas are not efficient.^7^ Data increasingly suggest that COVID-19 can be transmitted by airborne aerosols, marking the importance of particle filtration for preventing infections.^8,9,11^ In addition, viral transmission in hospital settings is associated with increased pathogen concentration in aerosolized particles (<5μm) inhaled by healthcare workers who are in direct contact with patients.^8^ Therefore, it is recommended that doctors and other healthcare professionals use N95 instead of surgical masks while performing high risk aerosol generating work such as intubation and dental work.^8,11^

The COVID-19 pandemic, particularly in the early months, overwhelmed the traditional supply chain creating a vital need for alternate PPE strategies.^,12^ It has become clear that hospitals needed to find an alternate infrastructure for the design and development of short-run manufacturing options of PPE.^13^ In addition, due to the fast spread of the virus and lack of information, many patient facing hospital staff were extremely worried about transmission in protocols that do not officially require the use of an N95. A clear example would be a nurse simply attending to a patient in a clinic who could have unknowingly contracted COVID-19 and therefore would be at risk of contracting it themselves as they only have the protection of a surgical mask and not an N95. Realizing this, the Innovation group at a medical cancer center set up a collaboration with the engineering school at a graduate research institution in order to develop and validate designs for alternate PPE. The goal was to come up with innovative designs that could be mass produced at a reasonable cost. The designs were also meant to be simple enough to be produced through community networks.

N95 respirators are more difficult and expensive to manufacture on a large scale compared to surgical masks, to an extent that some projects have explored ways to decontaminate and reuse N95 respirators.^19^ While surgical masks act as efficient barriers against viral droplets such as influenza, drug-resistant Tuberculosis, and COVID-19; they are associated with reduced aerosol filtration efficiency.^12,13,14^ Specifically, research shows that surgical masks reduce droplet exposure by 25 fold compared to only 2.8 fold in aerosol exposure.^15^ We hypothesize that this is largely due to the poor fit, prone to particles leakage, of surgical mask compared to N95 respirators. Specifically, areas around the cheeks, nasal sidewalls, and under the chin seem to be associated with poor fit (Fig.1A). 3D printing a complete facemask would require a significant amount of time for individual customization and material for printing. Furthermore, many materials used for 3D printing cannot be comfortably worn against the skin for long periods and cannot be sterilized. Given the drawbacks of manufacturing a complete mask, we aimed at creating an adjunct that would improve the face seal and increase the utility of a standard surgical mask.

**Figure 1:**
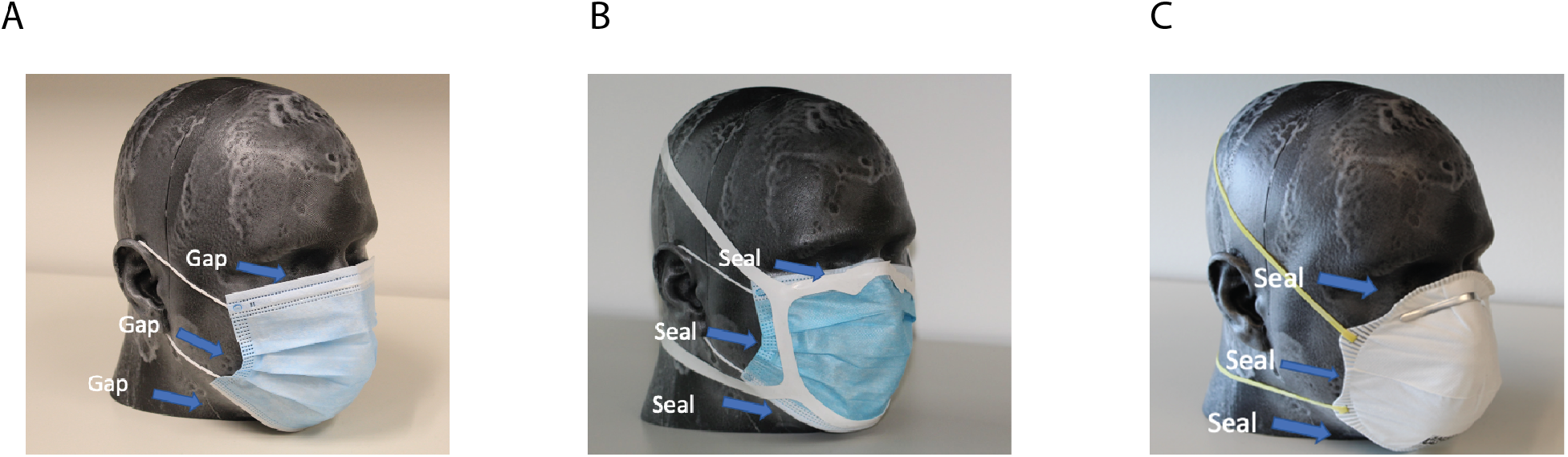
Improving the fit of a surgical mask using elastomeric harness. NIOSH 3D-printed large mannequin head wearing surgical mask alone (A), surgical mask with Harness Design 1 (B), and N-95 respirator (C)

A recent study shows that wearing a surgical mask with a rubber band alteration increases the fitted filtration efficiency (FFE), however, the work was only tested on one subject.^18^ Based on these designs a group has developed elastomeric harnesses, but there has been no data to show how particle filtration or fit are improved.^18^ Here, we show that different elastomeric harness designs cut from a single sheet of silicone rubber can improve the fit of surgical masks to the point that some individuals can pass the same fit tests used to evaluate N95 fit and particle leakage efficiency (Fig.1B). Central to developing a more aerosol-filtration efficient surgical mask lies the ability to successfully test it on a diverse group of people. In 2003 NIOSH conducted an anthropometric survey on 3997 respirator users stratified by gender, age, and race to inform fit tests procedures.^18^ Based on the results, they developed five face categories that represent most individuals. We used this information to improve our initial prototype design and then began to conduct OSHA approved tests on individuals with these harnesses.

Fit Tests measure the ambient microscopic particle concentration and the concentration of particles inside the mask. The ratio of these concentrations outputs the fit factor, which determines whether a surgical mask or an N95 respirator correctly fits the individual. First, we tested the harnesses on the five NIOSH-based face categories, then conducted a NIOSH approved fit test for quantitative analysis of aerosol filtration on 18 individuals wearing a level 3 surgical mask with a bendable wire around the nose. Finally, we repeated the fit test on 21 hospital staff wearing level 1 surgical masks without a bendable wire or adhesive features.

## MATERIALS AND METHODS

### Harness Designs Development

Both designs 1 and 2 were developed in the Illustrator application with the goal to improve the seal around 3 areas: nasal sidewalls, cheeks, and under the chin. Design 1 is relatively simple compared to design 2, which has increased material to loop around the nasal sidewalls to further reduce any aerosol transmission and/or inhalation (**Fig. 2**). Design 2.1 is similar to design 2 but with a shorter material to loop around the nasal sidewalls in order to reduce the protrusion of the mask under the eyes. The harnesses are cut from silicone material on the epilog laser cutting platform and disinfected before distributing to participants for fit testing.

**Figure 2:**
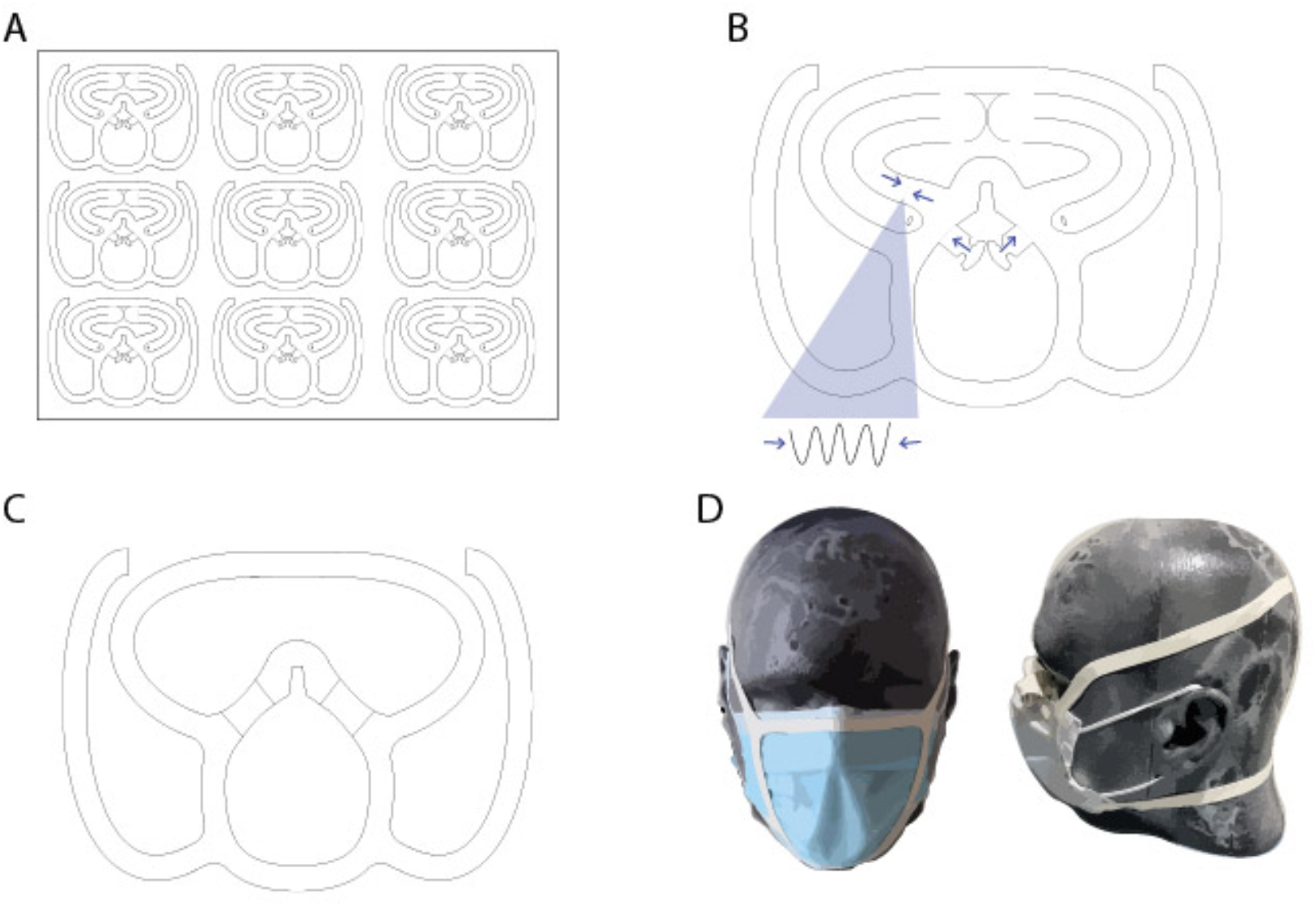
Harness designs production. Both harness designs 1, 2 and 2.1 were laser-cut using the epilog machine on a silicone material with a 1/32 inches in thickness (A). Harness Design 2 has more material in the nasal sidewalls area. The additional material is then folded multiple times (B) and secured around the nasal sidewalls (C). Once the material is secured, the harness is positioned over the surgical mask to improve the seal around the nose, cheeks and chin (D).

### Mannequin Head Testing Procedure

To test the effectiveness of the harness on the mannequin heads and examine potential “leak areas”, we used IR camera recordings. First, the mannequin heads wearing only a surgical mask were connected to a tube. On the opposite end of the tube, a temperature and pressure-controlled hair dryer was used to blow air inside the mannequin head, mimicking inhalation **(Fig.3)**. To show exhalation, the end of the hair dryer was connected to the opposite end of the tube, pulling back all the air that was previously introduced into the mannequin head. Video recordings using IR camera were taken for all mannequin head categories while wearing a surgical mask alone and with each harness design.

**Figure 3:**
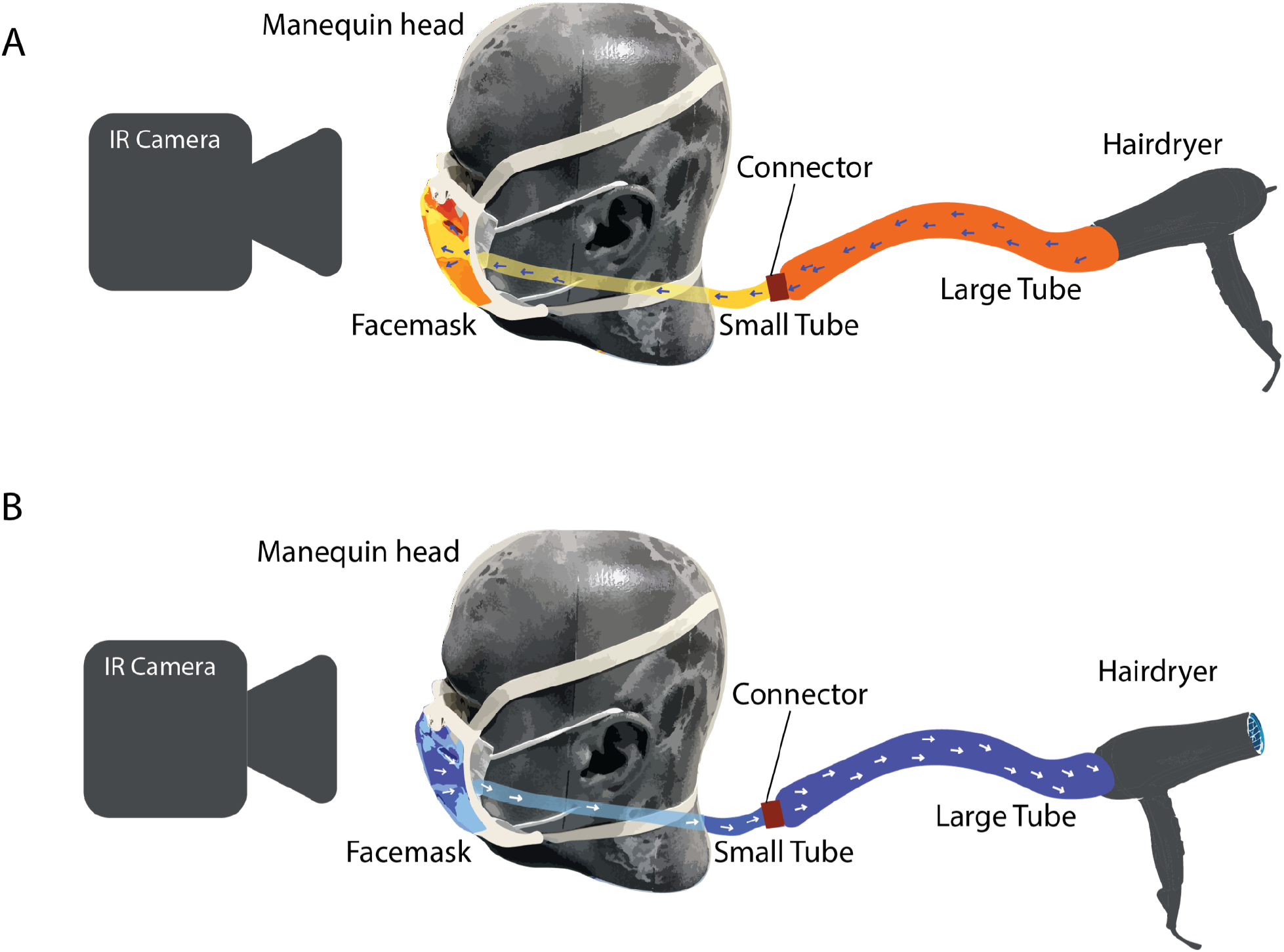
Mannequin Heads breathing simulation of exhalation (**A**) and Inhalation (**B**). Hairdryer introduces warm air into a large tube, which connects to a smaller tube directly in contact with the mannequin mouth. Warm air propagates inside the mask, and the seal efficiency is assessed using an IR camera. The protocol is performed for different mannequin sizes.

### Quantitative Fit Test procedure

Quantitative fit tests were conducted between October and December at a research institution and November and March at a medical cancer center. The Institutional Review Board at the research institution approved all the work done in this study and fit tests were performed considering the safety of participants by taking all COVID-19 prevention measures. The Environmental and Health Safety at the medical cancer conducted and approved all the fit tests done in this study. The TSI 8026 Particle Generator (MN) was used to supplement the ambient particle count in the room. Level 3 surgical masks with a bendable wire were used on all test subjects at the graduate research institution and level 1 regular surgical masks were used at medical cancer center. To connect the surgical mask to the probe, a testing port was installed on the surgical mask using the TSI probe kit. The subject attached the surgical mask to the probe by connecting the mask port and probe. Fit test measurements were recorded using the TSI 830 Portacount Pro (MN) producing a series of exercises as outlined in the OSHA quantitative test guidelines. In the first group, all participants did fit tests with designs 1 and 2 and select subjects did control fit tests with a surgical mask alone. At the medical center, all subjects did control fit tests with a surgical mask and fit tests on design 2.1. The OSHA quantitative fit tests take participants through multiple tests before producing a final fit test score. These multiple tests are normal breathing, deep breathing, head up and down, head side to side, talking, bending over and normal breathing again.

## RESULTS

In order to maximize both the face-seal as well as user-comfort, three different versions of the harness were tested. Design 1 significantly reduced the leak rate but failed to achieve reasonable face-seal along the nasal sidewalls. Design 2 improved face-seal with the addition of elastomeric material to increase the bulk in the region of the nasal sidewalls. While this extra material improved face-seal, it contributed to increased discomfort and blocked the user’s field of view during downward gaze. Consequently Design 2.1 was fabricated to include less material along the nasal sidewalls that was sufficient to maintain comfort and field of view, while also improving face-seal (**Fig. 2**)

Initially the harness design was tested on 3D printed mannequins representing the five NIOSH determined face categories.^18^ Inhalation and exhalation was simulated using a pressure-controlled hair dryer and an IR camera was used to detect the location of leaks (**Fig. 3**). IR imaging shows hotspots around the eyes and nasal sidewalls when the mannequin heads are only wearing the surgical masks with no harness, indicating significant air leakage in those areas (**Fig. 4**). Similar results were seen across all mannequin head sizes, with pronounced leaking on one side instead of both sides on smaller mannequin heads (**Fig. 4**.).

**Figure 4:**
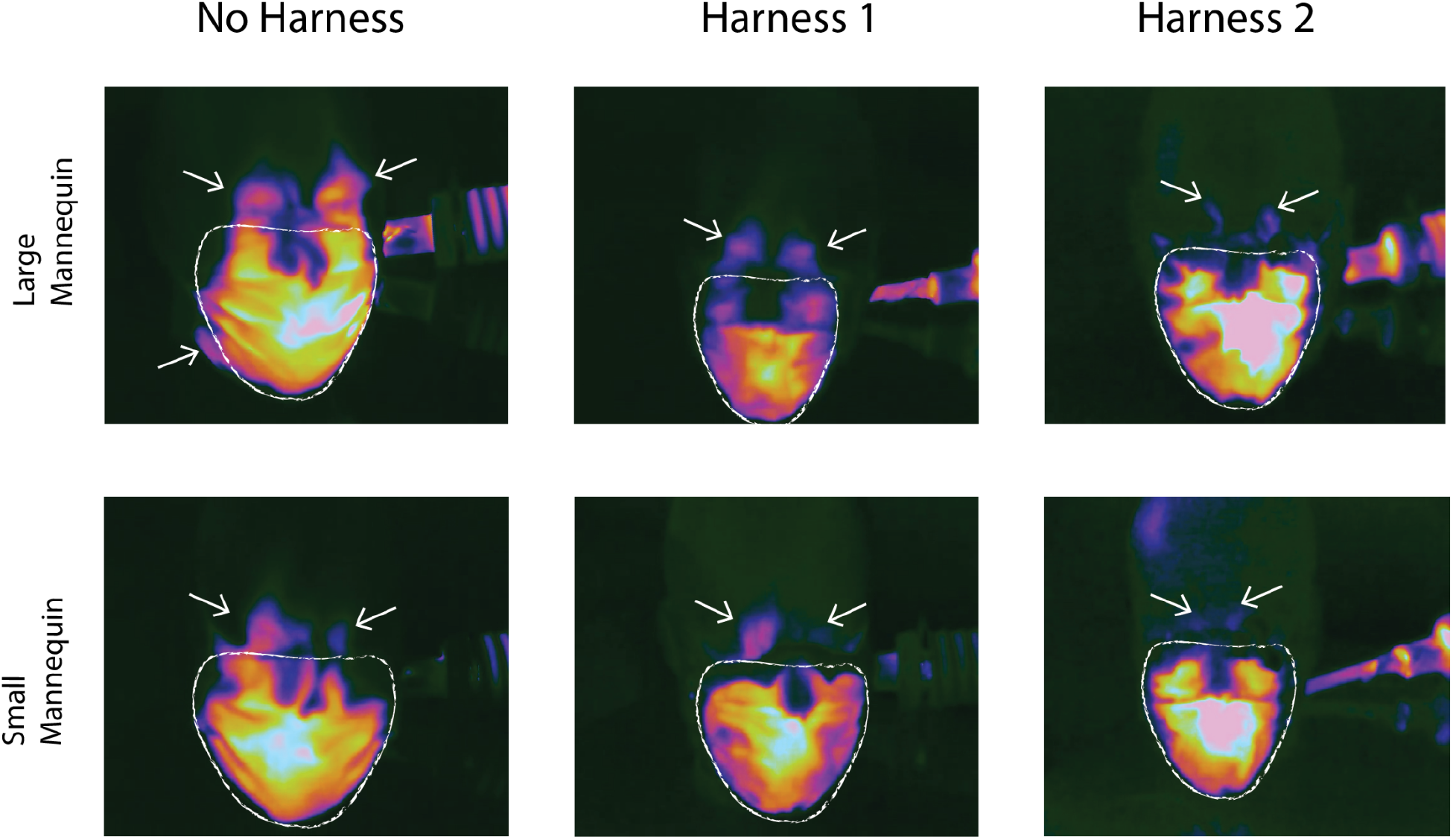
IR imaging of exhalation simulations in the large and small mannequin heads. (**A**) Mannequin heads wearing surgical mask only, (**B**) mannequin heads wearing surgical mask and harness design 1, (**C**) Mannequin heads wearing surgical mask and harness design 2.

Design 2, which includes more material along the nasal sidewalls, was noted to have reduced hot spots around the eyes and nasal sidewalls on IR imaging implying less leakage and a better fit. (**Fig. 4 and supplemental video 3**). These results were comparable to those obtained with a standard N95 respirator. In fact, due to the nature of this platform and the increased exhalation pressure from the hairdryer, N95 respirators also showed similar areas of leakage during IR imaging trials (**supplemental video 4**). Overall, in the context of this platform, harness Design 2 provided a significantly better face seal than Design 1 (**Fig. 4**).

OSHA approved quantitative fit tests using the TSI 830 Portacount Pro were used to evaluate the fit of both harness designs in 11 women and 7 men at a research institution. All 18 subjects had fit tests performed while wearing both designs 1 and 2. Control fit test measurements were taken on 11 participants while wearing a surgical mask without a harness. The passing score on the OSHA approved fit test is greater than or equal to 100. While wearing surgical masks alone the average Overall Fit Score was 3.21. None of the 18 individuals achieved a passing score (**Fig. 5A**). When fitted with design 1, 12 of the 18 individuals achieved a passing score of greater than 100 with an average Overall Fit Score of 120.83. When fitted with Design 2, 14 of the 18 individuals achieved a passing score with an average score of 152. All 12 of the individuals who achieved a passing score with design 1 also achieved a passing score when fitted with design 2 (**Fig. 5A)**. Seven of the 18 subjects passed the fit test with the maximum fit score of 200+ while wearing the second harness. As expected, when the two designs were tested on three male participants with beards, a passing score was not obtained, yet a marked increase in mask fit] was seen with design 2 compared to design 1 and surgical mask alone. Only one participant without facial hair failed the fit test with both designs 1 and 2.

**Figure 5:**
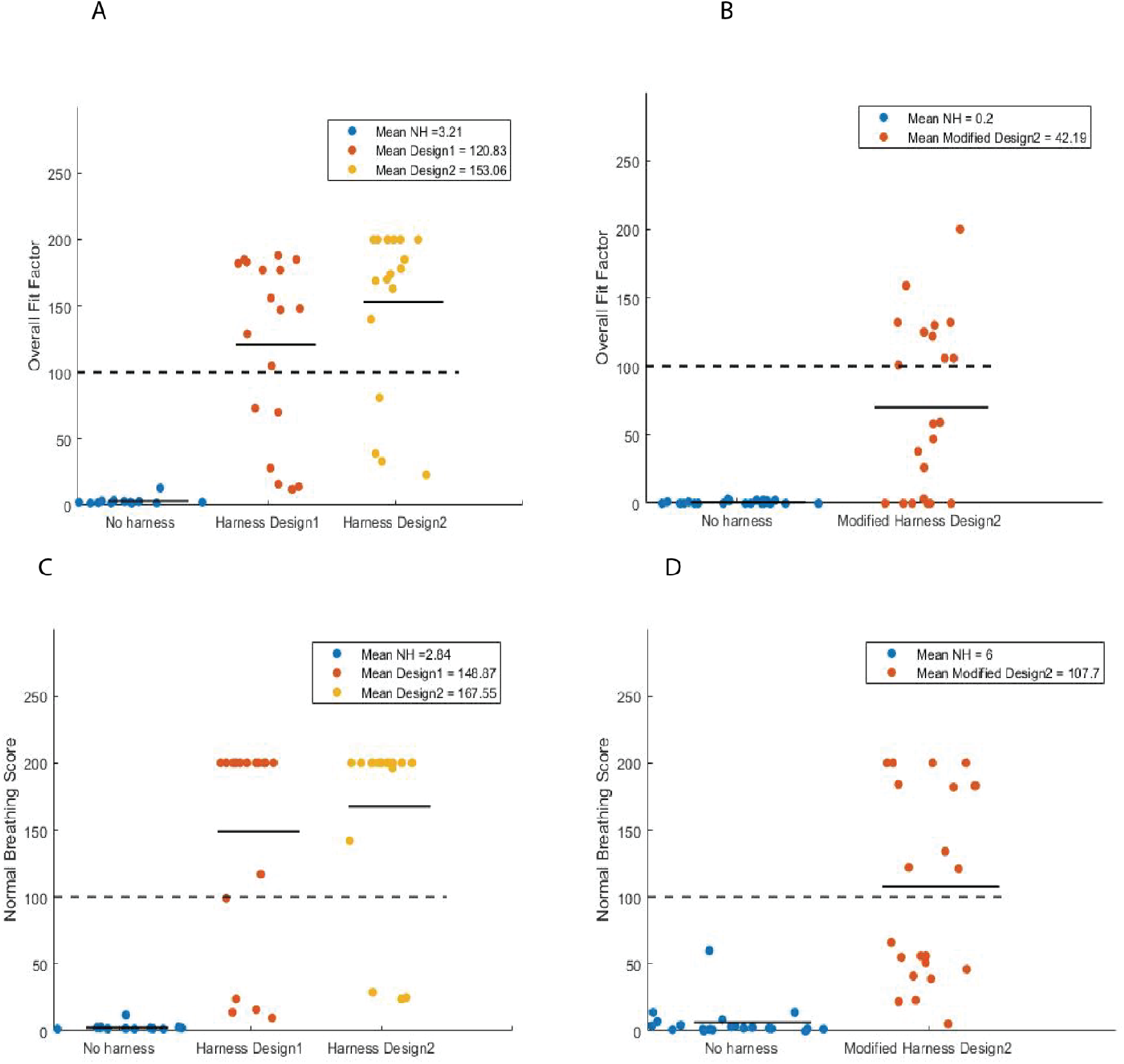
Elastomeric harnesses improve the seal of the surgical mask. (**A**) Overall fit test scores from individuals wearing harness designs 1 and 2. (**B**) Overall fit test scores of hospital staff wearing harness design 2.1. (**C**) Normal breathing test scores from individuals wearing harness designs 1 and 2. (**D**). Normal breathing scores of hospital staff wearing harness design 2.1.

Design 2.1 was tested on perioperative hospital staff at a medical cancer center. As mentioned earlier, Design 2.1 had been improved to be more comfortable and to cause less gaze obstruction. Fit tests were conducted on 10 women and 11 men. All 21 subjects had fit tests performed while wearing a surgical mask with and without the harness. Due to the features of the fit testing machine and protocol at the medical center, both Overall Fit Factor **(Fig 5A, 5B)** and Normal Breathing Score **(Fig 5C, 5D)** were measured. The fit test protocol used at medical center automatically generates a null value for the Overall Fit Factor if the subject obtains a quantitative failing score of less than 100. A Normal Breathing Score, on the other hand, is generated even if the subject fails the test with a score below 100. All participants received a quantitative value between 0 and 200+ for the Normal Breathing Score. All participants failed the fit test while wearing the surgical mask only **(Fig 5B)**. 10 participants passed the fit test while wearing the surgical mask and harness design 2.1 **(Fig 5B)**. All participants show a clear increase in their normal breathing score when wearing design 2.1 compared to the surgical mask alone. The average score without the harness was 6. The addition of the harness improved the average score almost 15-fold to 107.7 **(Fig 5D)**. All participants were then given two design 2.1 harnesses to wear around the hospital campus and daily life for the next 72 hours in order to provide qualitative data about the design and areas for improvement. A survey with questions pertaining to comfort and ease of use was sent to each of the participants. Of the 6 participants who completed the survey, 5 indicated that the harness was easier to use and more comfortable than an N95 respirator.

The mean normal breathing score of designs 1, 2 and 2.1 were 148.87, 167.55 and 107.7 respectively. These scores imply that design 2 has a better fit than design 1. Design 2.1 sacrifices some improvements in fit for better user comfort (**Fig. 5**).

## DISCUSSION

This study represents the successful collaboration between a medical center and engineering school to design and test a low cost, easy to manufacture, elastomeric harness that significantly improves the fit of a standard surgical mask. Infra-red imaging and OSHA approved fit testing was used to improve the design to optimize both fit and user comfort.

By studying the location of air leakage using IR imaging, we were able to determine that the nasal sidewalls region of the masks is the most prone to leakage when using our first elastomeric harness design (**supplemental videos 1 and 2**). In design 2, we see noticeable improvement of the harness fit on all mannequin heads by improving the seal around the nasal sidewalls (**Fig. 4 and supplemental video 3**). Surprisingly for some individuals the fit score decreased with the second design. We hypothesize that this is due to variations in facial features and comfort levels. This design improves the face seal and fit, but it caused increased discomfort in some participants, which prompted us to modify the design by reducing the amount of elastomeric material in the areas of discomfort.

While our data shows that both harness designs 1 and 2 perform better than harness design 2.1, care should be taken when making direct comparisons between tests performed at the medical center and those performed at the research institution. As previously discussed, the medical center testing protocol allocates an Overall Fit Score of 0 to everyone who doesn’t achieve a passing score of 100; however, the research institution protocol allocates a non-zero numerical Overall Fit Score even if the participant scores below 100. As such, it may not be accurate to compare these harness designs based on the average Overall Fit Scores alone. Another difference between these studies is the fact that the research institution used Level 3 medical surgical masks with a bendable wire and a particulate filtration efficiency (PFE) at 0.1 micron of 98%. The medical center team used level 1 non-adhesive surgical masks with a PFE of 95%. However, since all participants in both groups fail the fit test while wearing surgical mask, the nature of the masks alone is insufficient to achieve a passing fit score.

Design 2.1 was created to fit the daily tasks of perioperative hospital staff who occasionally require additional PPE such as eyeglasses and whose gaze is constantly downward when tending to patients in the operating room and clinic. Therefore, design 2.1 has less material folded along the nasal sidewalls which reduces gaze obstruction and increases comfort from the reduced pressure in this area. Considering the IR imaging from designs 1 and 2, we hypothesize that the lower score of design 2.1 is due to air leaking around the nasal sidewalls as there is not as tight a seal from the reduction of material.

## LIMITATIONS

The tradeoff between comfort and fit is of important consideration. While participants indicated that the first harness design can be worn comfortably for many hours, design 2 was reported to be relatively uncomfortable. Future work should explore other manufacturing techniques such as injection molding which can make the nasal sidewalls seal more comfortable and easier to assemble.

## CONCLUSIONS

Our work builds off of existing research on the efficacy of surgical masks and is intended to improve their aerosol filtration abilities. Past work clearly shows that N95 respirators are superior to surgical masks, especially in the context of aerosol transmission.^15^ We developed three low cost and re-usable harnesses that can be easily and cheaply manufactured on a large scale to improve the effectiveness of aerosol filtration of different types of surgical masks. While this work is especially relevant in the ongoing COVID-19 global pandemic, it provides further evidence for the need for improved fit of surgical masks in aerosol-generating procedures that are potentially infectious or present other biological harm.^8,11^As a collaboration between an engineering school and a medical center, this work gives insights on the different needs of the general public and health care workers. Future studies should consider the use of these harnesses with fabric masks associated with a higher or comparable filtration efficiency to the surgical mask.

## Data Availability

All data pertaining to this work, beyond what is shown here, is available upon request.

## ACKNOWLEDGEMENTS

We thank Dr. Guillaume Duret for the helpful discussions and MD Anderson Environmental Health and Safety Department. We thank Rice University’s COVID Fund for financial support of this multi-institutional project.

